# Fine particulate air pollution and neuropathology markers of Alzheimer’s disease in donors with and without APOE ε4 alleles – results from an autopsy cohort

**DOI:** 10.1101/2023.04.07.23288288

**Authors:** Grace M. Christensen, Zhenjiang Li, Donghai Liang, Stefanie Ebelt, Marla Gearing, Allan I. Levey, James J. Lah, Aliza P. Wingo, Thomas S. Wingo, Anke Huels

## Abstract

**Introduction:** Higher fine particulate matter (PM_2.5_) exposure has been found to be associated with Alzheimer’s disease (AD). PM_2.5_ has been hypothesized to cause inflammation and oxidative stress in the brain, contributing to neuropathology. A major genetic risk factor of AD, the apolipoprotein E (*APOE*) gene, has also been hypothesized to modify the association between PM_2.5_ and AD. However, little prior research exisits to support these hypotheses. Therefore, this paper aims to investigate the association between traffic-related PM_2.5_ and AD hallmark pathology, including effect modification by *APOE* genotype, in an autopsy cohort.

**Methods:** Brain tissue donors enrolled in the Emory Goizueta Alzheimer’s Disease Research Center (ADRC) who died before 2020 (n=224) were assessed for AD pathology including Braak Stage, Consortium to Establish a Registry for AD (CERAD) score, and the combined AD neuropathologic change (ABC score). Traffic-related PM_2.5_ concentrations were modeled for the metro-Atlanta area during 2002-2019 with a spatial resolution of 200-250m. One-, 3-, and 5-year average PM_2.5_ concentrations prior to death were matched to participants home address. We assessed the association between traffic-related PM_2.5_ and AD hallmark pathology, as well as effect modification by *APOE* genotype, using adjusted ordinal logistic regression models.

**Results:** Traffic-related PM_2.5_ was significantly associated with CERAD score for the 1-year exposure window (OR: 1.92; 95% CI: 1.12, 3.30), and the 3-year exposure window (OR: 1.87; 95%-CI: 1.01, 3.17). PM_2.5_ had harmful, but non-significant associations on Braak Stage and ABC score. The strongest associations between PM_2.5_ and neuropathology markers were among those without *APOE* ε*4* alleles (e.g., for CERAD and 1-year exposure window, OR: 2.31; 95% CI: 1.36, 3.94), though interaction between PM_2.5_ and *APOE* genotype was not statistically significant.

**Conclusions:** Our study found traffic-related PM_2.5_ exposure was associated with CERAD score in an autopsy cohort, contributing to epidemiologic evidence that PM_2.5_ affects Aβ deposition in the brain. This association was particularly strong among donors without *APOE* ε*4* alleles. Future studies should further investigate the biological mechanisms behind this assocation.

## Introduction

Dementia is a neurodegenerative condition with an increasing prevalence worldwide. It is estimated that by 2050, 131.5 million people worldwide will have dementia^1^. Neurodegeneration is caused by progressive neuronal breakdown in both the structure and function of the central nervous system (CNS)^2^. This degradation of the CNS is characterized by symptoms like memory loss, confusion, difficulty speaking, hallucinations, and paranoia^3^. Exposure to ambient and traffic-related air pollution, particularly particulate matter less than 2.5 micrometers in diameter (PM_2.5_), has been associated with dementia incidence and other measures of cognitive decline and dementia^4–6^. One review concluded that evidence suggests air pollution is causally associated with cognitive impairment based on the Bradford Hill guidelines for causality^4^. While these are interesting findings, the majority of these studies did not examine neuropathology or molecular endophenotypes that limit our ability to uncover potential molecular associations that underlie these associations.

Alzheimer’s disease (AD) is the most common cause of dementia, accounting for approximately 60-80% of dementia cases. The hallmark pathologies of AD are beta-amyloid (Aβ) plaques and neurofibrillary tangles (NFTs)^7^. Over time these changes associate with progressive brain injury and neurodegeneration^7^. AD has been found to be associated with ambient PM ^8,9^, as well as traffic-related PM_2.5_^10,11^. Traffic-related air pollutants are a major contributor to ambient air pollution levels, especially in urban environments. A hypothesized mechanism for the effect of PM_2.5_ on AD is through activation of microglia, resulting in neuroinflammation and increased intracellular reactive oxygen species (ROS), causing damage to brain tissue^2,12^. The majority of this evidence is from animal and *in vitro* models, as it is difficult to obtain the brain samples needed to assess these markers in human studies^2^.

To understand the relationship between AD and air pollution exposure, it is important to study human brain composition and pathology markers. Neuropathology markers like Braak stage, Consortium to Establish a Registry for AD (CERAD) score, and the combined AD neuropathologic change (ABC score) can provide information on severity of AD, but can only be obtained through autopsy. These markers represent the actual neuropathologic changes in the brain that are associated with AD and cognitive impairment^13^. Investigating hallmark neuropathologic changes, including NFTs and Aβ plagues, can help researchers understand the biological mechanisms underlying the progression of dementia and AD that incident cases or cognition tests cannot. Few autopsy cohorts have large enough sample sizes with personal information to investigate associations between these neuropathologies and environmental factors like air pollution. To our knowledge, only one study has investigated the association between PM_2.5_ and these neuropathology markers in humans. Researchers found inconclusive associations between Braak stage, CERAD score, ABC score and 10 year average PM_2.5_ concentration before death, although the vast majority of cases had low exposure levels in an autopsy cohort^14^. One study found an association between PM_2.5_ and CSF AD biomarkers in cognitively normal individuals, suggesting a link with AD brain pathology^15^.

Beyond environmental risk factors, there are also genetic risk factors for AD, the most notable of which is the ε4 variant of the apolipoprotein E (*APOE*) gene. It is hypothesized that certain isoforms of apolipoprotein E, specifically apoE4, can increase oxidative stress (presence of ROS) in the brain, and influence the development of AD^16^. Therefore, it is possible that the effect of PM_2.5_ on developing AD may be modified by *APOE* variant status. One prior study found that increased PM_2.5_ exposure in healthy adults was associated with faster rates of cognitive decline, with the impact greater in *APOE* ε*4* carriers^17^. However, another study using dementia diagnosis as the outcome did not find effect modification by *APOE* variant status^18^. There are few studies examining this effect modification on neuropathology markers in humans One study found no evidence of effect modification between *APOE* genotype and PM_2.5_ on Braak Stage, CERAD, or ABC score^14^. This gap in the literature could be due to the rarity of autopsy cohorts.

This study aims to address these critical gaps in the literature by leveraging a well-established autopsy cohort to investigate the effect of PM_2.5_ on AD hallmark pathology, as well as effect modification by *APOE* genotype.

## Methods

### Study population

Brain tissue donors were recruited by the Emory Goizueta Alzheimer’s Disease (AD) Research Center (ADRC). The ADRC maintains a brain bank to facilitate AD research. The majority of donors were research participants evaluated annually, and others werein the ADRC clinical core or patients treated by Emory physicians and diagnosed clinically with AD (biomarker defined) or probable AD. There were 1,011 donors in the brain bank by the third quarter of 2020. After applying the following inclusion criteria, 264 donors remained in the current analysis: 1) the availability of residential addresses within Georgia (GA) state; 2) age at death equal to or over 55 years; 3) deceased after 1999; 4) no missing values in outcomes (i.e., Braak Stage, CERAD, ABC) and key covariates including age at death, calendar year of death, race, sex, educational attainment, and *APOE* genotype. Written informed consent was provided for all donors. Samples were obtained using research protocols approved by the Emory University Institutional Review Board.

### Assessment of AD neuropathology

The ADRC performed thorough neuropathologic evaluations on the brains of all donors using established comprehensive research evaluations and diagnostic criteria^19^. All neuropathologic assessments included assessment of the severity of AD-related changes in neuropathology using a variety of stains and immunohistochemical preparations, as well as semi-quantitative scoring of multiple neuropathologic changes in numerous brain regions by experienced neuropathologists using published criteria. In this project, AD neuropathology was assessed using Braak stage, the CERAD, and ABC score. Braak stage is a staging scheme describing neurofibrillary tangles (NFTs) with six stages (Stage I-VI), and a higher stage indicates a wider distribution of NFTs in brain. CERAD score describes the prevalence of neuritic plaques with four levels from no neuritic plaques to frequent. ABC score combines the former two with the distribution of Amyloid plaques in the brain (Thal score) resulting in one of four levels of AD neuropathologic changes: not, low, intermediate, or high.

### Exposure assessment

Annual concentrations of traffic-related PM_2.5_ for 2002-2019 in the 20-county area of Metropolitan Atlanta, GA were estimated with a fine spatial resolution of 200-250m. Specifically, PM_2.5_ data for 2002-2011 were estimated for 250×250m grid cells evenly distributed throughout Metropolitan Atlanta; data for 2012-2019 had a spatial resolution of 200×200m. For 2002-2011, the Research LINE-source dispersion (R-LINE) model for near surface releases was applied for calculating annual averages and calibrated using a regression model approach with ten years of speciated PM_2.5_ measurements at three locations obtained via the receptor-based source apportionment Chemical Mass Balance Method with Gas Constraints (CMB-GC)^20,21^. For 2012-2019, we applied a land-use random forest model built on the training and test datasets comprised of the 2015 annual concentrations of traffic-related PM_2.5_ estimated by R-LINE and obtained from Atlanta Reginal Commission (ARC) ^22^, road inventory and traffic monitoring data shared by the Georgia Department of Transportation (GDOT)^23^, land cover data accessed via the National Land Cover Database^24^, and ambient PM_2.5_ data obtained from Atmospheric Composition Analysis^25^. The GDOT data data were based on actual measurements and contained accurate road geometry with traffic volume^26,27^. Four types of traffic input features were developed for highways and non-highways, respectively: 1) the shortest distance from the centroids of grids in ARC R-LINE data to the nearest local roads/highways, 2) the sum of road length in each grid, 3) the sum of annual average daily traffic (AADT) of roads in each grid, and 4) the AADT measured at traffic monitoring stations within or nearest to each grid. The land cover data had a spatial resolution of 30m with 15 categories of land use in GA. Eleven categories were generated including open water, barren, forest, shrub, herbaceous, agriculture, wetlands, and developed (open space, low intensity, medium intensity, and high intensity). The input features of proportions of land use for the 11 categories were calculated for each 200×200m grid^28^. We downloaded the annual ambient PM_2.5_ data with a spatial resolution of 0.01×0.01 degree and joined the ARC R-LINE grid cells with the ambient PM_2.5_ data spatially. Then, an ambient estimate for each grid was calculated as the input feature. In sum, the input features of 2015 derived from traffic and road inventory data, land use data, and ambient PM_2.5_ data entered the training of the air quality model, and the ARC traffic-related PM_2.5_ data of 2015 were used as the reference outoput. The random forest model was trained with the R package *randomForest*, and two user-defined parameters (i.e., the number of trees and the number of variables randomly tried at each split) were determined by a balance of the efficiency and the out-of-bag R^2^ value. Then, the final model was applied to the input features of 2012-2019 to predict annual traffic-related PM_2.5_. We spatially matched geocoded residential addresses prior to death to the centroid of closest grids and calculated the individual long-term exposures as the average of specific exposure windows: 1 year, 3 years, and 5 years prior to death.

### Confounder assessment

A directed acyclic graph (DAG) was created based on previous literature to select confounders and covariates for the association between PM_2.5_ and AD pathology (Figure S1). All models in the current analysis were adjusted for age at death, calendar year at death, race, sex, educational attainment, apolipoprotein E (*APOE*) genotype, and area deprivation index (ADI). Calendar year at death was adjusted for in addition to age at death to account for temporal changes in air pollution levels in the metro Atlanta area^29^. The race variable was binary as the current sample only contained White and Black participants. The educational attainment referred to the highest level of education that the subjects had completed, which was estimated based on number of years of education completed and classified into high school or less, college degree, and graduate degree. The *APOE* ε*4* allele is a well-known risk factor of developing AD, and the current analysis considered: 0, 1, and 2 ε*4* alleles. Also, a binary *APOE* genotype (ε4 absent vs. present) was used for testing the effect modification by the genotype. Binary *APOE* genotype was used for effect modification analyses to conserve statistical power in analyses (see Table 1 for a distribution of *APOE* ε*4* genotypes). The ADI is an indicator for neighborhood socioeconomic disadvantage at the census block group level assessed based on the theoretical domain of income, education, employment, and housing quality^30^. Our previous studies demonstrated that the association between long-term air pollution and cognitive function could be confounded by neighborhood-level socioeconomic status^31,32^.

**Table 1.**
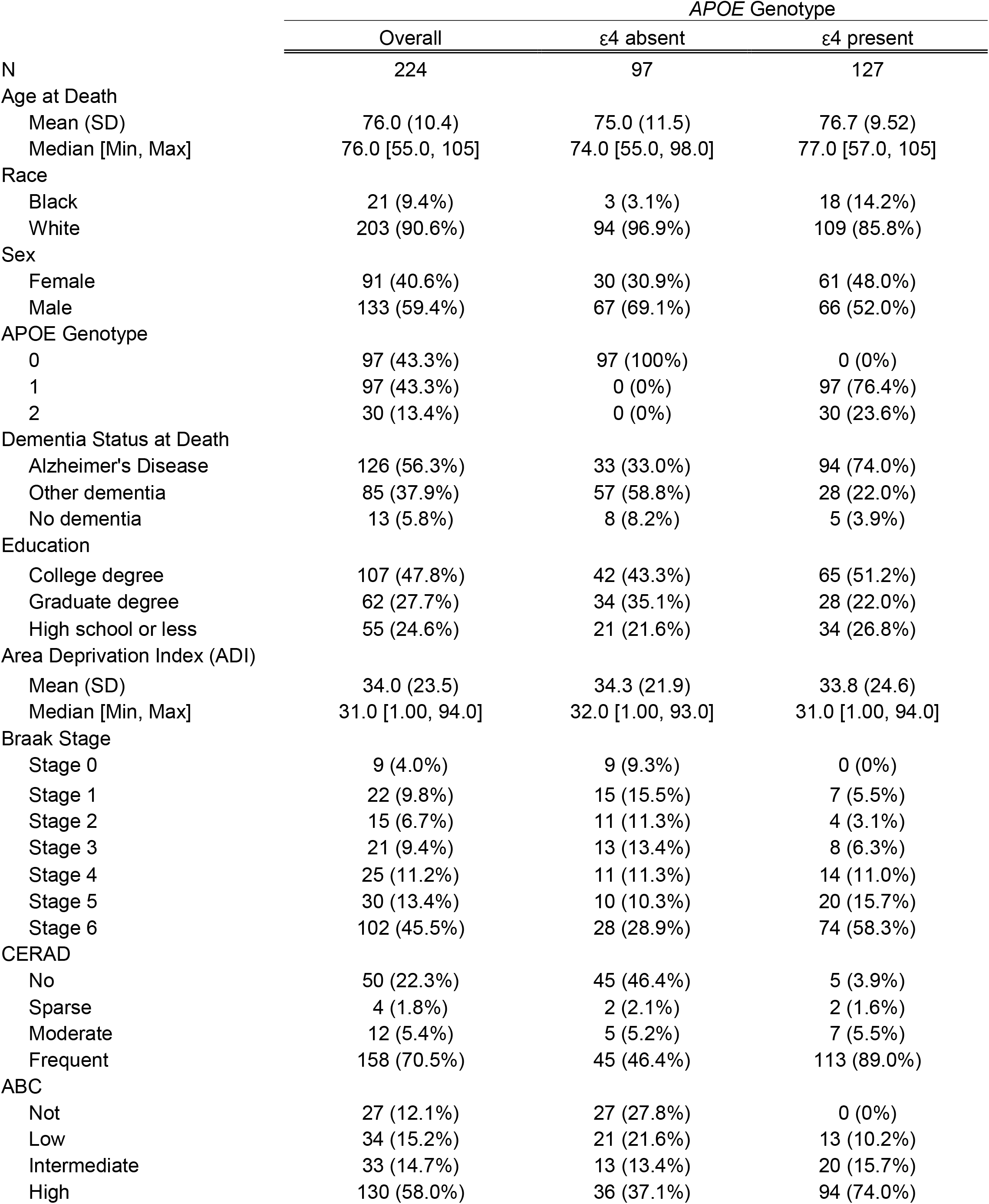
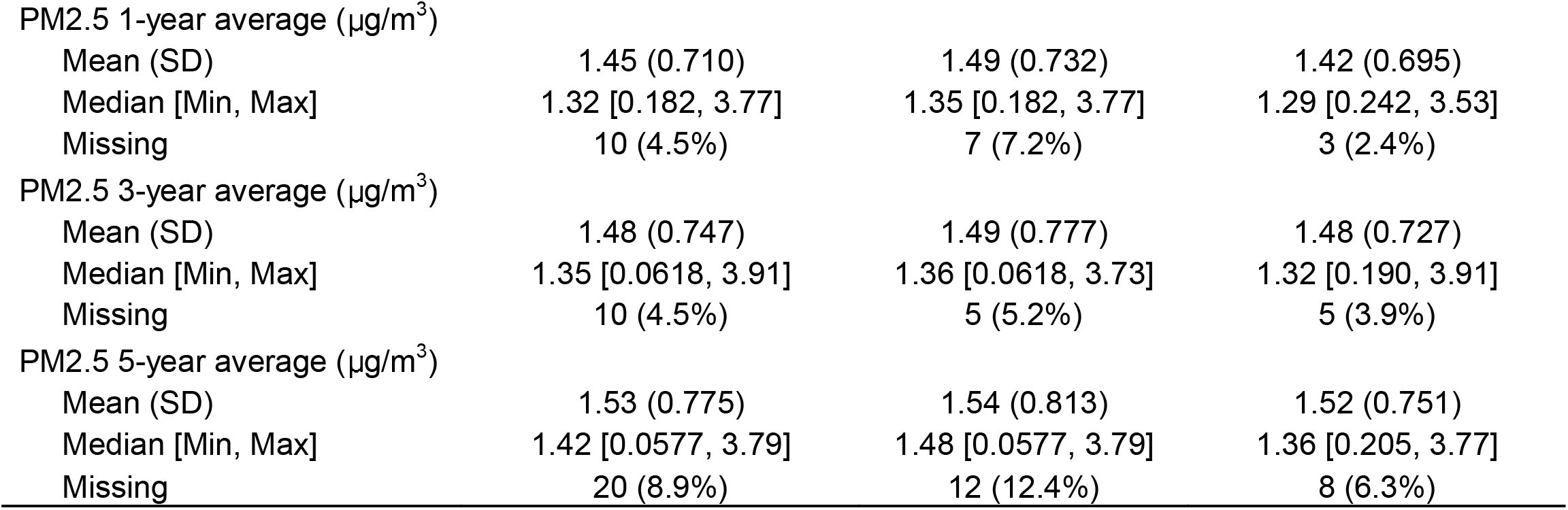
Descriptive statistics of study population stratified by *APOE* Genotype.

|  | *APOE* Genotype |  |  |
| --- | --- | --- | --- |
|  | Overall | ε4 absent | ε4 present |
| N | 224 | 97 | 127 |
| Age at Death |  |  |  |
| Mean (SD) | 76.0 (10.4) | 75.0 (11.5) | 76.7 (9.52) |
| Median [Min, Max] | 76.0 [55.0, 105] | 74.0 [55.0, 98.0] | 77.0 [57.0, 105] |
| Race |  |  |  |
| Black | 21 (9.4%) | 3 (3.1%) | 18 (14.2%) |
| White | 203 (90.6%) | 94 (96.9%) | 109 (85.8%) |
| Sex |  |  |  |
| Female | 91 (40.6%) | 30 (30.9%) | 61 (48.0%) |
| Male | 133 (59.4%) | 67 (69.1%) | 66 (52.0%) |
| *APOE* Genotype |  |  |  |
| 0 | 97 (43.3%) | 97 (100%) | 0 (0%) |
| 1 | 97 (43.3%) | 0 (0%) | 97 (76.4%) |
| 2 | 30 (13.4%) | 0 (0%) | 30 (23.6%) |
| Dementia Status at Death |  |  |  |
| Alzheimer's Disease | 126 (56.3%) | 33 (33.0%) | 94 (74.0%) |
| Other dementia | 85 (37.9%) | 57 (58.8%) | 28 (22.0%) |
| No dementia | 13 (5.8%) | 8 (8.2%) | 5 (3.9%) |
| Education |  |  |  |
| College degree | 107 (47.8%) | 42 (43.3%) | 65 (51.2%) |
| Graduate degree | 62 (27.7%) | 34 (35.1%) | 28 (22.0%) |
| High school or less | 55 (24.6%) | 21 (21.6%) | 34 (26.8%) |
| Area Deprivation Index (ADI) |  |  |  |
| Mean (SD) | 34.0 (23.5) | 34.3 (21.9) | 33.8 (24.6) |
| Median [Min, Max] | 31.0 [1.00, 94.0] | 32.0 [1.00, 93.0] | 31.0 [1.00, 94.0] |
| Braak Stage |  |  |  |
| Stage 0 | 9 (4.0%) | 9 (9.3%) | 0 (0%) |
| Stage 1 | 22 (9.8%) | 15 (15.5%) | 7 (5.5%) |
| Stage 2 | 15 (6.7%) | 11 (11.3%) | 4 (3.1%) |
| Stage 3 | 21 (9.4%) | 13 (13.4%) | 8 (6.3%) |
| Stage 4 | 25 (11.2%) | 11 (11.3%) | 14 (11.0%) |
| Stage 5 | 30 (13.4%) | 10 (10.3%) | 20 (15.7%) |
| Stage 6 | 102 (45.5%) | 28 (28.9%) | 74 (58.3%) |
| CERAD |  |  |  |
| No | 50 (22.3%) | 45 (46.4%) | 5 (3.9%) |
| Sparse | 4 (1.8%) | 2 (2.1%) | 2 (1.6%) |
| Moderate | 12 (5.4%) | 5 (5.2%) | 7 (5.5%) |
| Frequent | 158 (70.5%) | 45 (46.4%) | 113 (89.0%) |
| ABC |  |  |  |
| Not | 27 (12.1%) | 27 (27.8%) | 0 (0%) |
| Low | 34 (15.2%) | 21 (21.6%) | 13 (10.2%) |
| Intermediate | 33 (14.7%) | 13 (13.4%) | 20 (15.7%) |
| High | 130 (58.0%) | 36 (37.1%) | 94 (74.0%) |

|  |  |  |  |  |
| --- | --- | --- | --- | --- |
| PM2.5 1-year average ($\mu\text{g}/\text{m}^3$) | | | | |
| Mean (SD) | 1.45 (0.710) | 1.49 (0.732) | 1.42 (0.695) |  |
| Median [Min, Max] | 1.32 [0.182, 3.77] | 1.35 [0.182, 3.77] | 1.29 [0.242, 3.53] |  |
| Missing | 10 (4.5%) | 7 (7.2%) | 3 (2.4%) |  |
| PM2.5 3-year average ($\mu\text{g}/\text{m}^3$) | | | | |
| Mean (SD) | 1.48 (0.747) | 1.49 (0.777) | 1.48 (0.727) |  |
| Median [Min, Max] | 1.35 [0.0618, 3.91] | 1.36 [0.0618, 3.73] | 1.32 [0.190, 3.91] |  |
| Missing | 10 (4.5%) | 5 (5.2%) | 5 (3.9%) |  |
| PM2.5 5-year average ($\mu\text{g}/\text{m}^3$) | | | | |
| Mean (SD) | 1.53 (0.775) | 1.54 (0.813) | 1.52 (0.751) |  |
| Median [Min, Max] | 1.42 [0.0577, 3.79] | 1.48 [0.0577, 3.79] | 1.36 [0.205, 3.77] |  |
| Missing | 20 (8.9%) | 12 (12.4%) | 8 (6.3%) |  |

### Statistical analysis

The association between PM_2.5_ exposures and measured neuropathology outcomes was evaluated by ordinal logistic regression, considering the neuropathology markers as ordered categorical variables. The effect estimates for PM_2.5_ on neuropathology markers were reported as odds ratios associated with a one-unit (µg/m^3^) increase in PM_2.5_, 95% confidence intervals (CI) were calculated using the delta method. For the assessment of effect modification by *APOE* genotype, we added a product term in the model between dichotomous *APOE* genotype (binary) and PM_2.5_ exposures. The effect estimates for PM_2.5_ among ε4 carriers and non-carriers were reported respectively, and the 95% confidence intervals were calculated. All models were adjusted for confounders as described above. For power, dichotomous *APOE* genotype was used in main analyses, a sensitivity analysis using a three level *APOE* genotype variable (0, 1, or 2 ApoE ε4 alleles) was also conducted to evaluate a potential dose-response relationship.

Several sensitivity analyses were conducted to assess the robustness of results. First, we replaced ordinal logistic regression with multiple linear regression which considered the neuropathology markers as continuous variables. Next, we replaced ordinal logistic regression with traditional (binary) logistic regression analyses, for which neuropathology outcomes were categorized as the following: CERAD (0-1/negative vs 2-3/positive); Braak Stage (0-2/negative vs 3-6/positive); ABC (0-1/negative vs 2-3/positive). Finally, to investigate if effect modification with *APOE* genotype differs by race, an ordinal logistic regression analysis was conducted with White participants only. There were too few black participants to conduct a separate analysis for Black ancestry only. All analyses were conducted in R (version 4.2.0).

## Results

### Study population

Mean age at death for participants included in this analysis was 76.0 years (SD: 10.4 years). Participants were majority white (90.6%), majority male (59.4%) and were well educated (75.5% with a college degree or higher). In our sample, 43.3% had one *APOE* ε*4* copy, and 13.4% had two copies. Over half of participants in our sample were diagnosed with AD before death (56.3%), and 37.9% were diagnosed with other dementias. Almost half of participants were classified as having the highest Braak stage (45.5%), and over half were in the highest CERAD (70.5%) and ABC (58.0%) categories. (Table 1, Table S1). Concentrations of traffic-related PM_2.5_ in Metro Atlanta have decreased over the last decades^27,29^, therefore median traffic-related PM_2.5_ level increased as the exposure window was lengthened. For example, the 1-year average median concentration across participants was 1.32 μg/m^3^ (IQR: 0.18, 3.77), 3-year average median concentration was 1.35 μg/m^3^ (IQR: 0.06, 3.91), and the 5-year average median concentration was 1.42 μg/m^3^ (IQR: 0.06, 3.79) (Table 1, Figure S2). The 224 participants included in our analysis had similar sociodemographic characteristics and *APOE* genotype distribution compared to the full ADRC autopsy cohort, although participants included in this analysis were significantly older, and had significantly higher Braak stage, CERAD score, and ABC score compared to the original ADRC cohort (Table S1).

### Traffic-related PM_2.5_ exposure and neuropathology markers

In ordinal logistic regression models adjusted for age at death, calendar year at death, race, sex, education, *APOE* genotype, and ADI, PM_2.5_ showed a harmful association with all neuropathology metrics at all exposure windows. PM_2.5_ was significantly associated with CERAD score in models using a 1-year exposure window (OR: 1.92; 95% CI: 1.12, 3.30), and a 3-year exposure window (OR: 1.87; 95%-CI: 1.01, 3.17) (Figure 1, Table S2). PM_2.5_ also showed a harmful association with Braak stage and ABC score, but the association was not statistically significant at any exposure window (Figure 1, Table S2).

**Figure 1.**
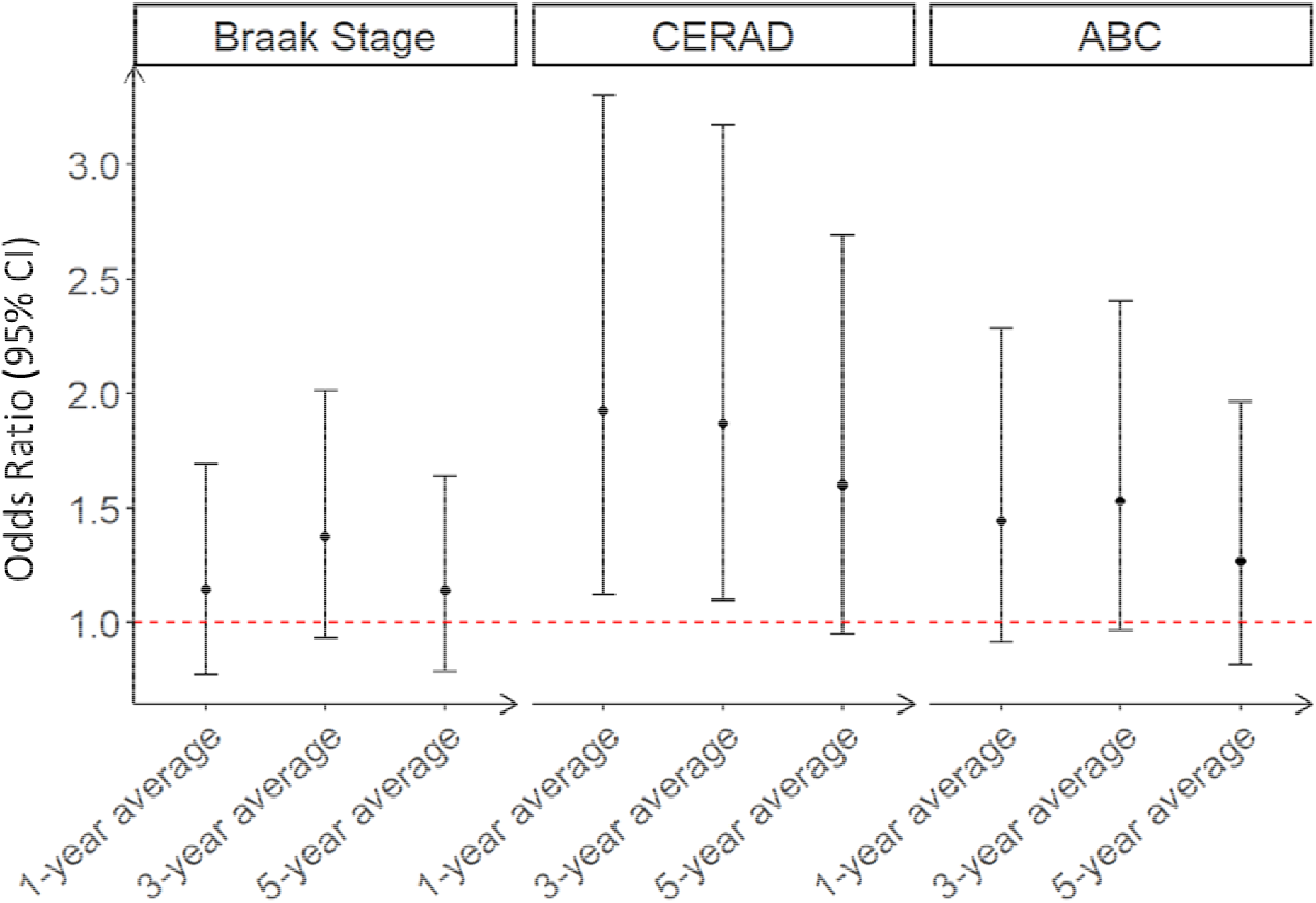
Associations between PM_2.5_ exposure windows and neuropathology markers. Associations shown are odds ratios and 95% CIs of ordinal logistic regression models for the effect of PM_2.5_ on neuropathology markers. Models adjusted for age at death, calendar year at death, race, sex, *APOE* status, education, and ADI.

Associations between PM_2.5_ and all neuropathology markers were stronger among those without the *APOE* ε*4* variant. While the interaction was not statistically significant, there was a consistent pattern suggesting effect modification. Among participants without the *APOE* ε*4* allele, PM_2.5_ was significantly harmful at all time windows on all neuropathology markers (Figure 2, Table S3). The largest effect estimates were associated with CERAD score (1-year exposure window (OR: 2.31; 95% CI: 1.36, 3.94); 3-year exposure window (OR: 2.31; 95% CI: 1.25, 3.61); 5-year exposure window (OR: 1.80; 95% CI: 1.07, 3.00)). PM_2.5_ was not significantly associated with any neuropathology marker among those with the *APOE* ε*4* allele (Figure 2, Table S3).

**Figure 2.**
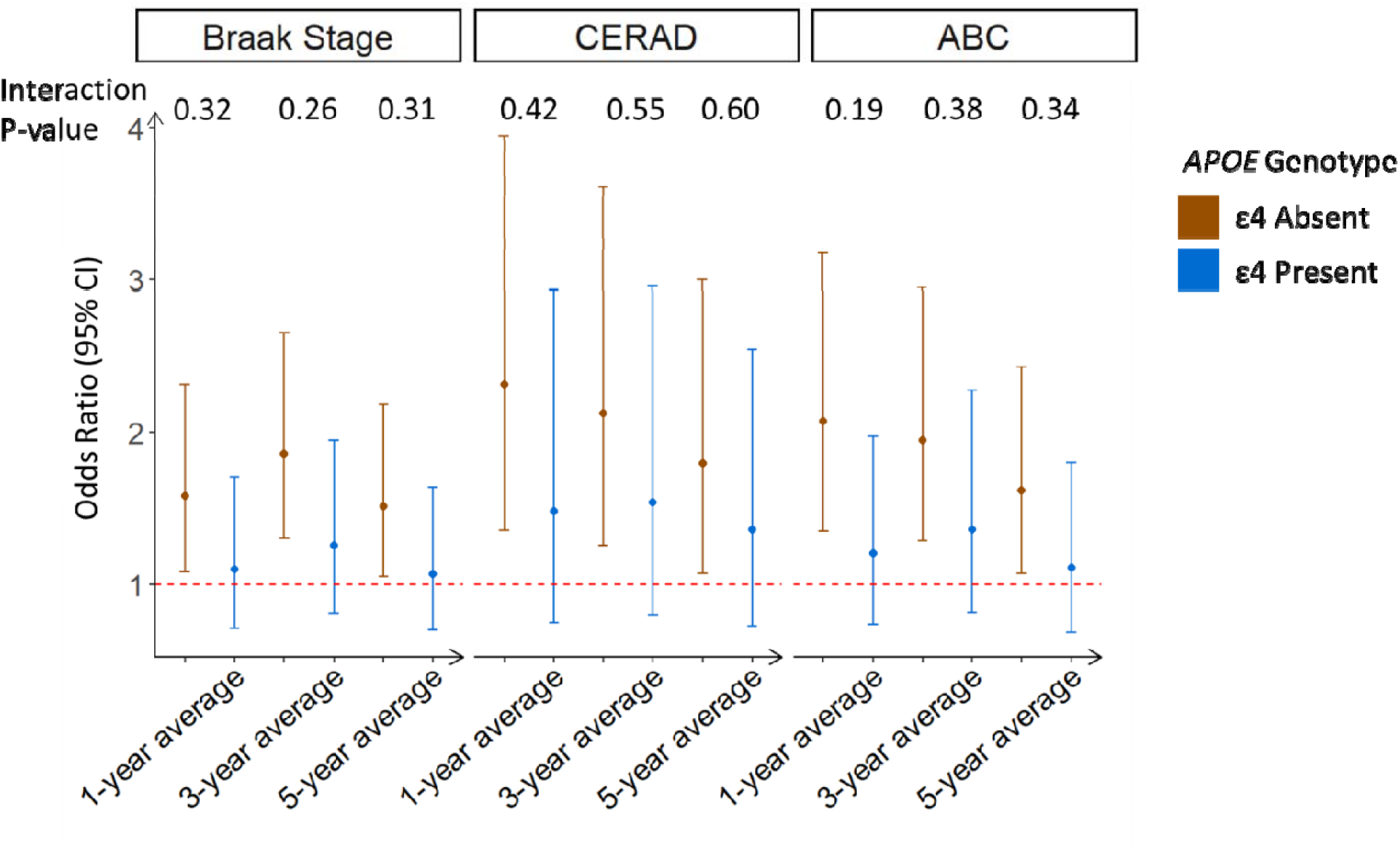
Association between PM_2.5_ and neuropathology markers stratified by *APOE* genotype (ε4 absent vs. present). Associations were estimated using ordinal logistic regression, models adjusted for age at death, calendar year at death, race, sex, education, ADI, and a PM_2.5_\**APOE* genotype interaction term.

### Sensitivity analyses

Though not statistically significant, there were harmful associations between PM_2.5_ and neuropathology markers in all linear regression models (table S4, figure S3), which is in line with the results from the ordinal logistic regression analysis. In all adjusted linear regression models investigating interaction by *APOE* ε*4* allele, the effect of PM_2.5_ was strongest among those without the *APOE* ε*4* allele. Among those without the *APOE* ε*4* allele, PM_2.5_ was significantly associated with Braak stage at the 3-year average exposure window and with CERAD score for all exposure windows. For the ABC score, 1- and 3-year PM_2.5_ exposure windows were significantly associated with ABC score in those without the *APOE* ε*4* allele. Among those with the *APOE* ε*4* allele, PM_2.5_ was not significantly associated with any neuropathology markers. Similar to the ordinal logistic regression analysis, the interaction term was not statistically significant in any model (Figure S4, Table S5). Logistic regression analyses also showed similar patterns as orinal logistic regression analyses, though the binary neuropathology outcomes were underpowered to identify statistically significant associations (Table S6, Table S7). Ordindal logistic regression analyses with White participants only had similar results as the main orindal logistic regression analyses (Figure S5).

Ordinal logistic regression analysis with interaction by a three level APOE genotype (0, 1, 2 ε4 alleles) found similar effects of PM_2.5_ on neuropathology markers as the dichotomous APOE genotype analyses. Associations between PM_2.5_ and the neuropathology markers Braak stage and ABC score were strongest among those with 0 copies of the ApoE ε4 allele (Figure S6). For the CERAD score, the effect of PM_2.5_ was similar between those with 0 and 2 copies of ApoE ε4, but associations among carriers of 2 copies of ApoE ε4 had the widest confidence interval due to the small samples size (n=30). Again, the interaction term was not statistically significant in any model.

## Discussion

In this study of 224 brain tissue donors from the ADRC in Metro Atlanta, we observed statistically significant harmful associations between traffic-related PM_2.5_ and the CERAD score, a semi-quantitative rating of neuritic plaques, as well as similar but non-significant harmful associations with Braak stage and ABC score. Associations between PM_2.5_ and neuropathology markers were stronger among those without any copies of the *APOE* ε*4* allele, though the interaction terms were not statistically significant.

There is a growing body of mechanistic and epidemiologic evidence linking air pollution exposure with AD and related dementias^5,10,33^. Most previous studies have focused on links between air pollution and cognitive function or incident cognitive impairment or dementia^4,6^. Incident dementia is often studied using diagnostic codes on insurance billing claims and medical records, facilitating studies with large sample sizes; however, billing data are known to miss true dementia cases and provide no information on the degree of AD pathophysiology^34^. The only other study investigating PM_2.5_ and neuropathology markers in autopsy samples found non-significant harmful associations between PM_2.5_ and CERAD but not with the ABC score or Braak stage^14^. While their association with CERAD was not statistically significant, the direction of association was in line with our study, in which we also found the strongest associations for CERAD. Our study differs in three main aspects from the previous study by Shaffer et al.: 1) their sample size was larger than ours (832 versus 224 donors), 2) their study was population-based, a major difference, which could have negatively influenced their statistical power to detect associations, and 3) the PM_2.5_ concentrations were measured as the 10-year ambient average, as opposed to our 1-, 3-, and 5-year traffic-related averages. In their autopsy cohort from Seattle, WA, the 10-year average (SD) PM_2.5_ exposure was 8.2 (1.9) μg/m^3^. In contrast, our 1-year average (SD) traffic-related PM_2.5_ exposure was 1.46 (0.71) μg/m^3^. The lower average PM_2.5_ exposure in our cohort is due to the restriction to traffic-related PM_2.5_, which is a subgroup of ambient PM_2.5_ exposure that was used in Shaffer et al. We decided to use traffic-related PM_2.5_ as opposed to ambient because our study population is in an urban area, where traffic-related air pollutants are a major source source of ambient pollution^11^. Additionally, traffic-related pollutants can be more easily targeted for interventions as the source is known. The major advantage of our exposure modelling approach is the high spatial resolution of 200-250m, which allowed us to identify finer-scaled spatial variations in urban exposure concentrations than with traditional modelling approaches, and consequently increased the statistical power to detect associations with neuropathology markers.

Our study as well as the study by Shaffer et al.^14^ found that PM_2.5_ was most strongly associated with CERAD score. CERAD neuropathology score examines neuritic plaque density^35^. These plaques are deposits of the Aβ protein in the grey matter of the brain, surrounded by degenerating neurites. Inflammation is a risk factor for Aβ plaques, and neuritic plaques have been shown to contain activated microglia and other signs of an inflammatory response^36,37^. Ambient and traffic-related PM_2.5_, and other air pollutants, are known to contribute to neuroinflammation^12^. While not directly comparable to our autopsy studies, Aβ plaques have been consistently associated with air pollution exposure in other studies examining Aβ via positron emission tomography (PET) scans, cerebrospinal fluid (CSF) or plasma. Studies in cognitively normal or mildly impaired populations found 5-year average PM_10_ ^38^ and annual average NO_2_^15^ to be associated with higher Aβ deposits in their brain measured via PET scan. Another study found similar associations in a population composed of participants with either MCI or dementia^39^, a population that is more comparable to our own autopsy cohort, in which 71.9% were diagnosed with either AD or other forms of dementia. Limited research has explored the association between air pollution and Aβ measured in CSF or plasma, but two recent studies showed an association between PM_2.5_ absorbance and CSF AD biomarkers for higher levels of brain Aβ deposition^15^ as well as between PM_2.5_ exposure and plasma-based measures of Aβ^40^. Therefore, our study makes an important contribution to a growing body of evidence that PM_2.5_ affects Aβ deposition and neuritic plaque formation in the brain. Understanding the mechanism by which air pollution impacts brain health remains an important endeavor, because it is an important piece of information for the determination of causality.

We found stronger associations between PM_2.5_ and all neuropathology markers among those without an *APOE* ε*4* allele, though the interaction term was not statistically significant. *APOE* ε4 is the strongest known risk factor for AD, and may be masking the weaker effects of PM_2.5_ on Aβ deposition in the brain. In our sample, 56.4% of participants had at least 1 copy of the ε4 allele, which may have masked some of the effect of PM_2.5_ on neuropathology markers in the model that only adjusted for *APOE* genotype as a confounder. Overall, the epidemiologic evidence for effect modification by *APOE* genotype has been mixed^14,15,17,18^. In line with our findings, analyses using AD biomarkers such as centiloid values (CL)^15^, a measure of brain Aβ deposition, and AD incidence^18^ found stronger associations with PM_2.5_ among non-carriers of *APOE* ε*4*. In contrast, analyses using CSF NfL levels as the outcome^15^, a biomarker of subcortical axonal degeneration, or cognitive decline^17^, PM_2.5_ showed a stronger association among *APOE* ε*4 carriers*. However, both studies were conducted in participants without cognitive impairment^15^ or at least with a lower prevalence of dementia (∼11%)^17^, compared to our autopsy cohort. These mixed results between studies investigating effect modification by *APOE* genotype may be due to heterogeneity in outcome measurement and age of participants. Comparing effect modification by *APOE* genotype between studies using AD incidence, neuropsychological tests for cognitive decline, and various biomarkers may not yield similar results as the underlying role of *APOE* may be different among different cognitive outcomes. As more studies investigating AD are published, the relationship between PM_2.5_, *APOE* genotype, and AD will become more clear.

One limitation of this study is that the ADRC cohort is enriched with cases with AD and other dementias, meaning the brain bank is a convenience sample and not a population-based sample. Typically, brain banks are not created with epidemiology studies in mind and are focused on disease cases. The abundance of AD cases in this sample decreased the variability of neuropathology markers, especially among the *APOE* ε*4* carriers. Additionally, this study has a relatively small sample size. Though, few studies have this large of an autopsy sample, which is necessary for measuring neuropathology markers, and is a major strength of this paper. The ADRC is composed of primarily White and highly educated participants of high socioeconomic status. This may make the results not generalizable to other populations. Another limitation is that the only residential address available to connect to air pollution measurements was the address at death. This may lead to misclassification of the exposure in the larger time windows, and explains the attenuation towards the null of effect estimates for the 3- and 5-year exposure windows.

## Conclusion

Our study found traffic-related PM_2.5_ exposure was associated with CERAD score at autopsy, contributing to a growing body of evidence that PM_2.5_ affects Aβ deposition in the brain. This association was particularly strong among donors without *APOE* ε*4* alleles, suggesting that *APOE* ε*4*, which is the strongest known risk factor for AD, could mask the (weaker) effects of PM_2.5_ on Aβ deposition in the brain. More research is needed to establish causality for the association between PM_2.5_ and AD, including epidemiologic and mechanistic studies.

## Data Availability

All data produced in the present study are available upon reasonable request to the authors

## Funding Sources and Acknowledgements

This work was supported by the HERCULES Pilot Project via NIEHS P30ES019776 (Huels), the Goizueta Alzheimer’s Disease Research Center: Pilot Grant via NIA P50AG025688 (Huels/Liang), the Rollins School of Public Health Dean’s Pilot and Innovation Grant (Huels) and NIA R01AG079170 (Huels/Wingo). GMC was supported by the NIEHS T32 Training Program in Environmental Health and Toxicology (5T32ES12870). The air pollution exposure assessment was supported by the NIH grant R21ES032117 (Liang). We also want to thank Dr. Jeremy A. Sarnat (Emory), Dr. Armistead Russell (Georgia Tech), Ms. Kyung-Hwa Kim and Ms. Abby Marinelli from the Atlanta Regional Commission for providing data and guidance towards the air pollution exposure assessment.

