## Supplemental Materials for "Fine particulate air pollution and neuropathology markers of Alzheimer’s disease in donors with and without APOE ε4 alleles – results from an autopsy cohort"

**Table S1.** Comparison of descriptive statistics between ARDC Brain Bank participants included in the study and the rest of the cohort.

|  | Included in Study | |  |
| --- | --- | --- | --- |
|  | No | Yes | P-value |
| N | 536 | 224 |  |
| Age at Death |  |  |  |
| Mean (SD) | 74.1 (10.1) | 76.0 (10.4) | 0.0197 |
| Median [Min, Max] | 74.0 [55.0, 98.0] | 76.0 [55.0, 105] |  |
| Race |  |  |  |
| Black | 31 (5.8%) | 21 (9.4%) | 0.206 |
| Hawaiian | 3 (0.6%) | 0 (0%) |  |
| White | 448 (83.6%) | 203 (90.6%) |  |
| Missing | 54 (10.1%) | 0 (0%) |  |
| Sex |  |  |  |
| Female | 256 (47.8%) | 91 (40.6%) | 0.079 |
| Male | 280 (52.2%) | 133 (59.4%) |  |
| APOE genotype |  |  |  |
| 0 | 213 (39.7%) | 97 (43.3%) | 0.469 |
| 1 | 194 (36.2%) | 97 (43.3%) |  |
| 2 | 48 (9.0%) | 30 (13.4%) |  |
| Missing | 81 (15.1%) | 0 (0%) |  |
| Dementia Status at Death |  |  |  |
| AD | 306 (57.1%) | 123 (54.9%) | 0.585 |
| No dementia | 18 (3.4%) | 4 (1.8%) |  |
| Other dementia | 103 (19.2%) | 38 (17.0%) |  |
| Missing | 109 (20.3%) | 59 (26.3%) |  |
| Education |  |  |  |
| College degree | 60 (11.2%) | 107 (47.8%) | 0.899 |
| Graduate degree | 35 (6.5%) | 62 (27.7%) |  |
| High school or less | 35 (6.5%) | 55 (24.6%) |  |
| Missing | 406 (75.7%) | 0 (0%) |  |
| Braak Stage |  |  |  |
| Stage 0 | 42 (7.8%) | 9 (4.0%) | <0.001 |
| Stage 1 | 61 (11.4%) | 22 (9.8%) |  |
| Stage 2 | 82 (15.3%) | 15 (6.7%) |  |
| Stage 3 | 40 (7.5%) | 21 (9.4%) |  |
| Stage 4 | 42 (7.8%) | 25 (11.2%) |  |
| Stage 5 | 109 (20.3%) | 30 (13.4%) |  |
| Stage 6 | 160 (29.9%) | 102 (45.5%) |  |
| CERAD |  |  |  |
| No | 147 (27.4%) | 50 (22.3%) | 0.027 |
| Sparse | 22 (4.1%) | 4 (1.8%) |  |
| Moderate | 47 (8.8%) | 12 (5.4%) |  |
| Frequent | 320 (59.7%) | 158 (70.5%) |  |
| ABC |  |  |  |
| Not | 84 (15.7%) | 27 (12.1%) | 0.068 |
| Low | 117 (21.8%) | 34 (15.2%) |  |
| Intermediate | 71 (13.2%) | 33 (14.7%) |  |
| High | 264 (49.3%) | 130 (58.0%) |  |

**Table S2.** Effect estimates (95% CIs) of ordinal logistic regression models for the effect of PM_2.5_ on neuropathology markers. Models adjusted for age at death, calendar year at death, race, sex, APOE status, education, and ADI.

|  | Braak Stage | CERAD | ABC |
| --- | --- | --- | --- |
| 1-year average | 1.15 (0.77, 1.69) | **1.92 (1.12, 3.30)** | 1.45 (0.92, 2.28) |
| 3-year average | 1.37 (0.94, 2.02) | **1.87 (1.01, 3.17)** | 1.53 (0.94, 2.40) |
| 5-year average | 1.14 (0.79, 1.64) | 1.60 (0.95, 2.70) | 1.27 (0.82, 1.97) |

**Table S3.** Effect estimates (95% CIs) of ordinal logistic regression models with interaction between *APOE* genotype and PM_2.5_, for the effect of PM_2.5_ on neuropathology markers stratified by *APOE* genotype. Models adjusted for age at death, calendar year at death, race, sex, *APOE* status, education, and ADI.

|  | *APOE* Genotype | |  |
| --- | --- | --- | --- |
|  | ε4 Absent | ε4 Present | Interaction P-value |
| **Braak Stage** | | | |
| 1-year average | **1.58 (1.08, 2.31)** | 1.10 (0.71, 1.71) | 0.32 |
| 3-year average | **1.86 (1.31, 2.65)** | 1.26 (0.81, 1.95) | 0.26 |
| 5-year average | **1.51 (1.05, 2.18)** | 1.07 (0.70, 1.64) | 0.38 |
| **CERAD** | | | |
| 1-year average | **2.31 (1.36, 3.94)** | 1.48 (0.75, 2.93) | 0.42 |
| 3-year average | **2.13 (1.25, 3.61)** | 1.54 (0.80, 2.96) | 0.55 |
| 5-year average | **1.80 (1.07, 3.00)** | 1.36 (0.73, 2.55) | 0.60 |
| **ABC** | | | |
| 1-year average | **2.07 (1.35, 3.18)** | 1.20 (0.76, 1.97) | 0.19 |
| 3-year average | **1.95 (1.29, 2.95)** | 1.36 (0.81, 2.27) | 0.38 |
| 5-year average | **1.62 (1.07, 2.43)** | 1.11 (0.69, 1.80) | 0.34 |

**Table S4.** Effect estimates (95% CIs) of linear regression models for the effect of PM_2.5_ on neuropathology markers. Models adjusted for age at death, calendar year at death, race, sex, *APOE* status, education, and ADI.

|  | **Braak Stage** | **CERAD** | **ABC** |
| --- | --- | --- | --- |
| 1-year average | 0.17 (-0.25, 0.59) | 0.17 (-0.09, 0.44) | 0.14 (-0.09, 0.37) |
| 3-year average | 0.30 (-0.07, 0.67) | 0.17 (-0.7, 0.41) | 0.17 (-0.4, 0.37) |
| 5-year average | 0.16 (-0.20, 0.52) | 0.13 (-0.10, 0.36) | 0.09 (-0.11, 0.29) |

**Table S5.** Effect estimates (95% CIs) of linear regression models with interaction between *APOE* genotype and PM_2.5_, for the effect of PM_2.5_ on neuropathology markers stratified by *APOE* genotype. Models adjusted for age at death, calendar year at death, race, sex, *APOE* status, education, and ADI.

|  | *APOE* Genotype | |  |
| --- | --- | --- | --- |
|  | ε4 Absent | ε4 Present | Interaction P-value |
| **Braak Stage** | | | |
| 1-year average | 0.44 (-0.12, 1.01) | 0.16 (-0.34, 0.67) | 0.4 |
| 3-year average | **0.57 (0.07, 1.06)** | 0.26 (-0.20, 0.72) | 0.32 |
| 5-year average | 0.39 (-0.11, 0.88) | 0.12 (-0.34, 0.58) | 0.4 |
| **CERAD** | | | |
| 1-year average | **0.46 (0.11, 0.81)** | 0.13 (-0.18, 0.44) | 0.11 |
| 3-year average | **0.42 (0.10, 0.73)** | 0.11 (-0.18, 0.40) | 0.12 |
| 5-year average | **0.34 (0.03, 0.65)** | 0.09 (-0.20, 0.37) | 0.2 |
| **ABC** | | | |
| 1-year average | **0.34 (0.03, 0.64)** | 0.12 (-0.16, 0.39) | 0.22 |
| 3-year average | **0.34 (0.07, 0.62)** | 0.14 (-0.12, 0.39) | 0.23 |
| 5-year average | 0.25 (-0.16, 0.53) | 0.06 (-0.19, 0.31) | 0.27 |

**Table S6.** Effect estimates (95% CIs) of logistic regression models for the effect of PM_2.5_ on neuropathology markers. Binary neuropathology outcomes were categorized as the following: CERAD (0-1/negative vs 2-3/positive); Braak Stage (0-2/negative vs 3-6/positive); ABC (0-1/negative vs 2-3/positive). Models adjusted for age at death, calendar year at death, race, sex, *APOE* status, education, and ADI.

|  | Braak Stage | CERAD | ABC |
| --- | --- | --- | --- |
| 1-year average | 1.35 (0.63, 2.86) | 1.94 (0.90, 4.16) | 1.75 (0.87, 3.54) |
| 3-year average | 1.57 (0.75, 3.32) | 1.82 (0.91, 4.63) | 1.70 (0.88, 3.28) |
| 5-year average | 1.41 (0.71, 2.78) | 1.56 (0.82, 2.99) | 1.48 (0.80, 2.74) |

**Table S6.** Effect estimates (95% CIs) of logistic regression models with interaction between *APOE* genotype and PM_2.5_, for the effect of PM_2.5_ on neuropathology markers stratified by *APOE* genotype. Binary neuropathology outcomes were categorized as the following: CERAD (0-1/negative vs 2-3/positive); Braak Stage (0-2/negative vs 3-6/positive); ABC (0-1/negative vs 2-3/positive). Models adjusted for age at death, calendar year at death, race, sex, *APOE* status, education, and ADI.

|  | APOE Genotype | |  |
| --- | --- | --- | --- |
|  | Absent | Present | Interaction P-value |
| Braak Stage | | | |
| 1-year average | 1.69 (0.70, 4.11) | 0.86 (0.29, 2.54) | 0.28 |
| 3-year average | **1.66 (0.71, 3.86)** | 1.37 (0.42, 4.50) | 0.78 |
| 5-year average | 1.43 (0.66, 3.07) | 1.36 (0.45, 4.07) | 0.94 |
| CERAD | | | |
| 1-year average | 2.18 (0.94, 5.05) | 1.29 (0.94, 5.05) | 0.45 |
| 3-year average | 2.07 (0.96, 4.48) | 1.17 (0.36, 3.80) | 0.39 |
| 5-year average | 1.74 (0.85, 3.54) | 1.09 (0.35, 3.33) | 0.46 |
| ABC | | | |
| 1-year average | **2.4 (1.03, 5.62)** | 1.01 (0.38, 2.64) | 0.13 |
| 3-year average | 1.97 (0.92, 4.25) | 1.21 (0.45, 3.28) | 0.40 |
| 5-year average | 1.66 (0.82, 3.37) | 1.15 (0.46, 2.92) | 0.50 |

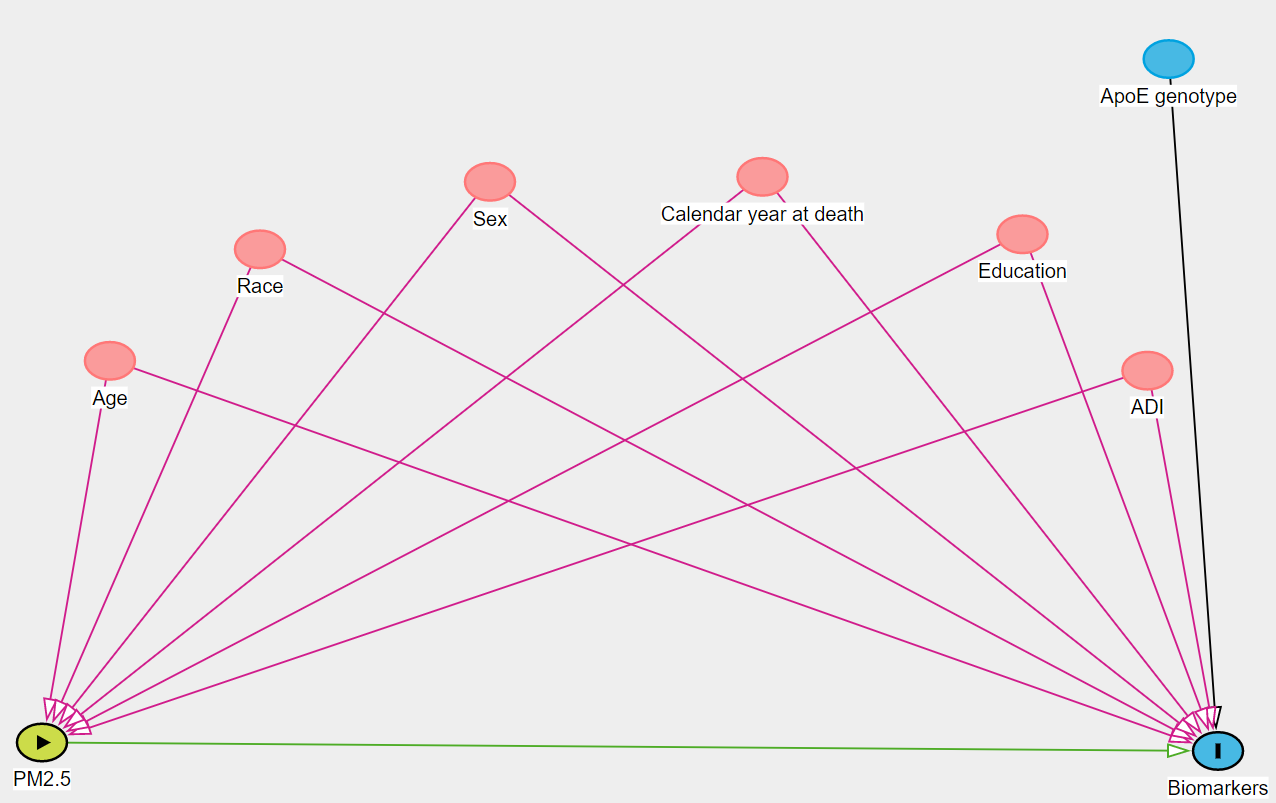

**Figure S1.** Directed Acyclic Graph (DAG) showing the relationships between PM_2.5_, neuropathology markers, and covariates.

A.	B.

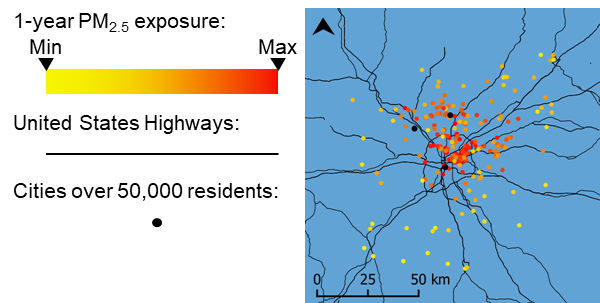

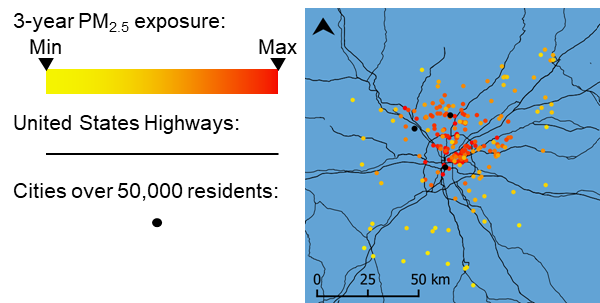

C.

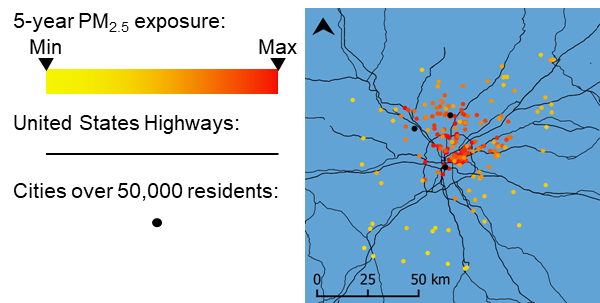

**Figure S2.** Air pollution exposure of ADRC participants in Metro-Atlanta. A. 1-year exposure window. B. 3-year exposure window. C. 5-year exposure window.

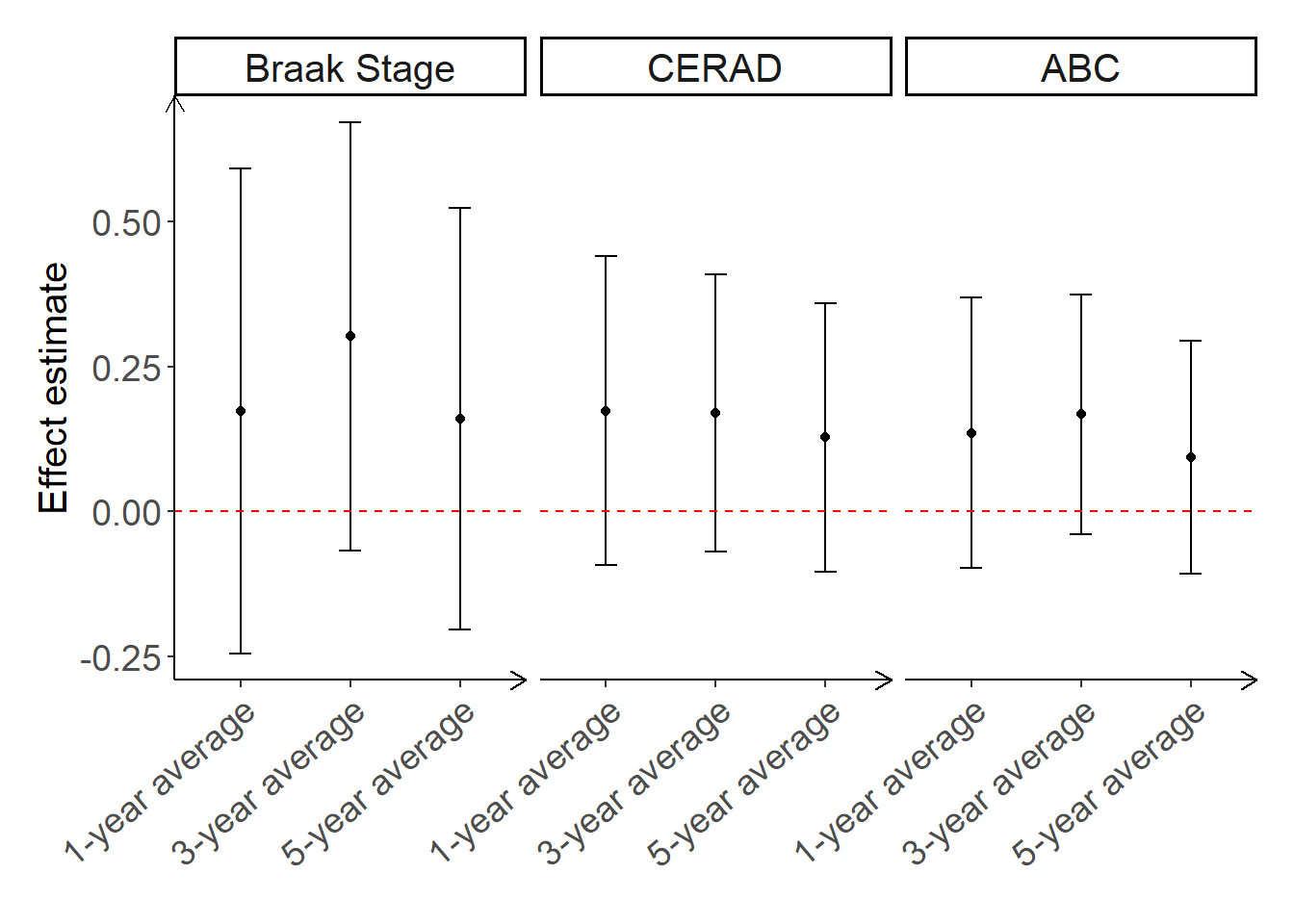

**Figure S3.** Associations between PM_2.5_ exposure windows and neuropathology markers. Associations shown are beta estimates and 95% CIs of adjusted linear regression models for the effect of PM_2.5_ on neuropathology markers. Models adjusted for age at death, calendar year at death, race, sex, *APOE* status, education, and ADI.

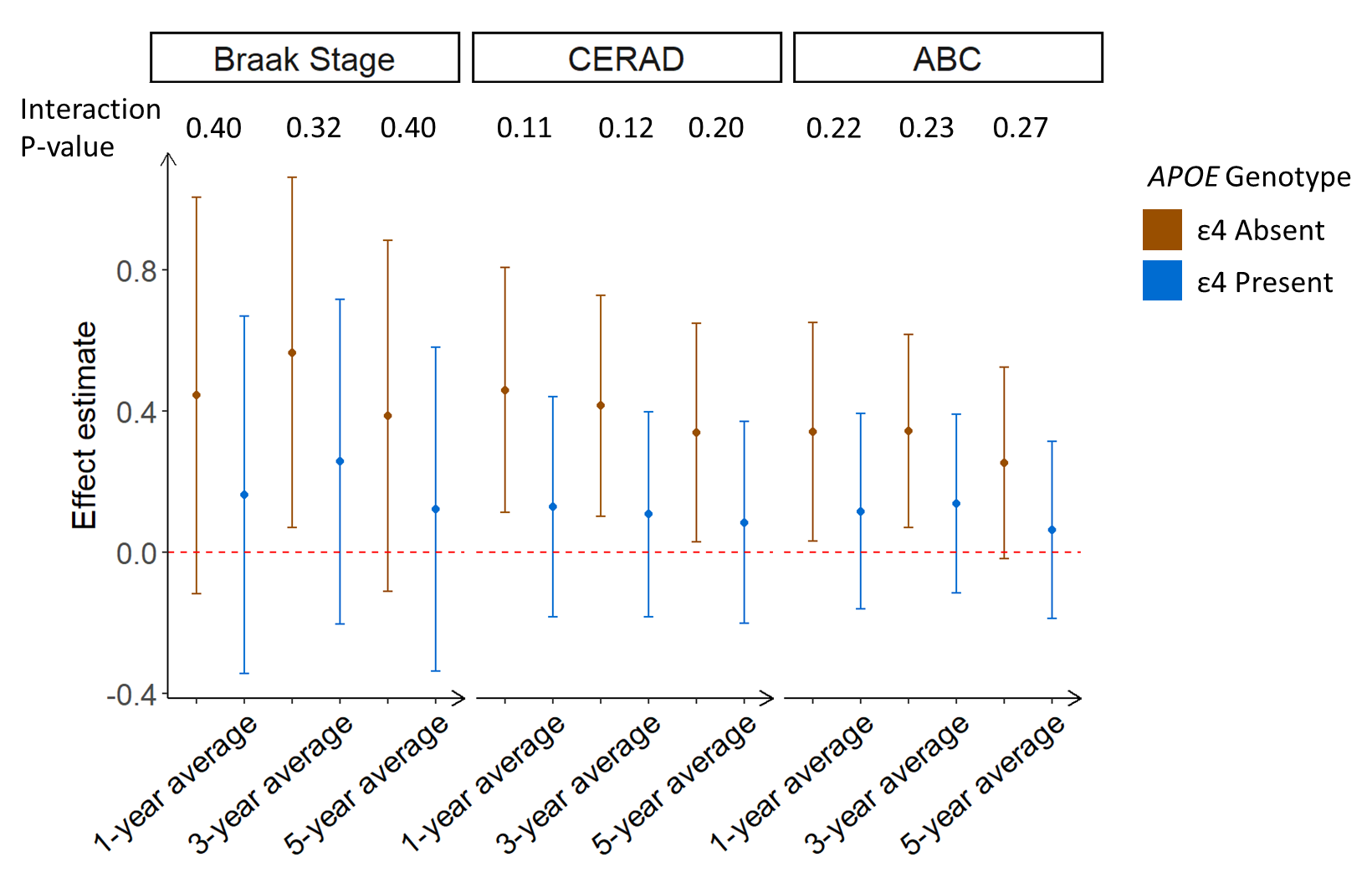

**Figure S4.** Association between PM_2.5_ and neuropathology markers stratified by *APOE* genotype (ε4 absent vs. present). Associations were estimated using adjusted linear regression, models adjusted for age at death, calendar year at death, race, sex, education, ADI, and a PM_2.5_**APOE* genotype interaction term.

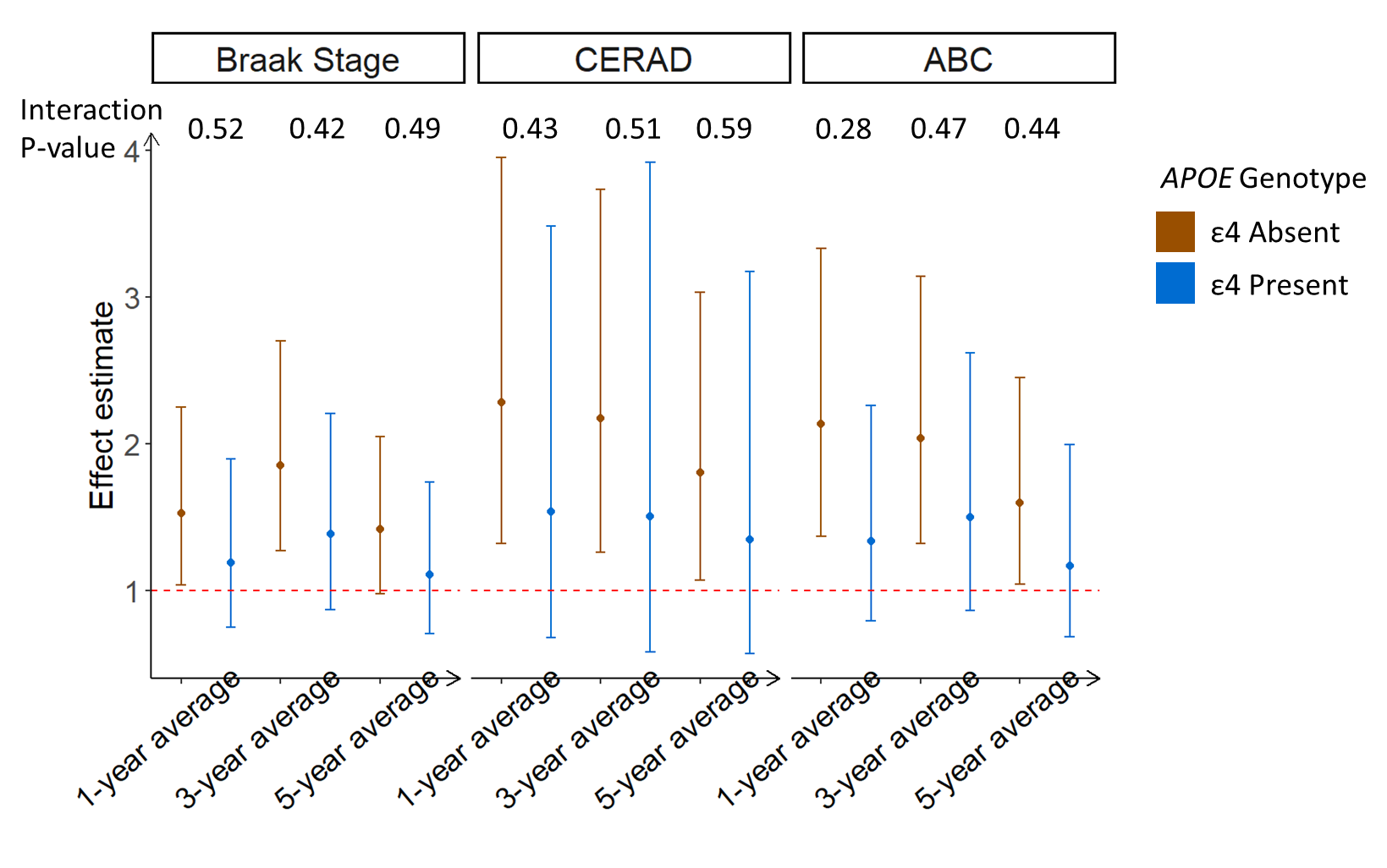

**Figure S5.** Association between PM_2.5_ and neuropathology markers stratified by *APOE* genotype (ε4 absent vs. present) among white participants only. Associations were estimated using ordinal logistic regression, models adjusted for age at death, calendar year at death, race, sex, education, ADI, and a PM_2.5_**APOE* genotype interaction term.

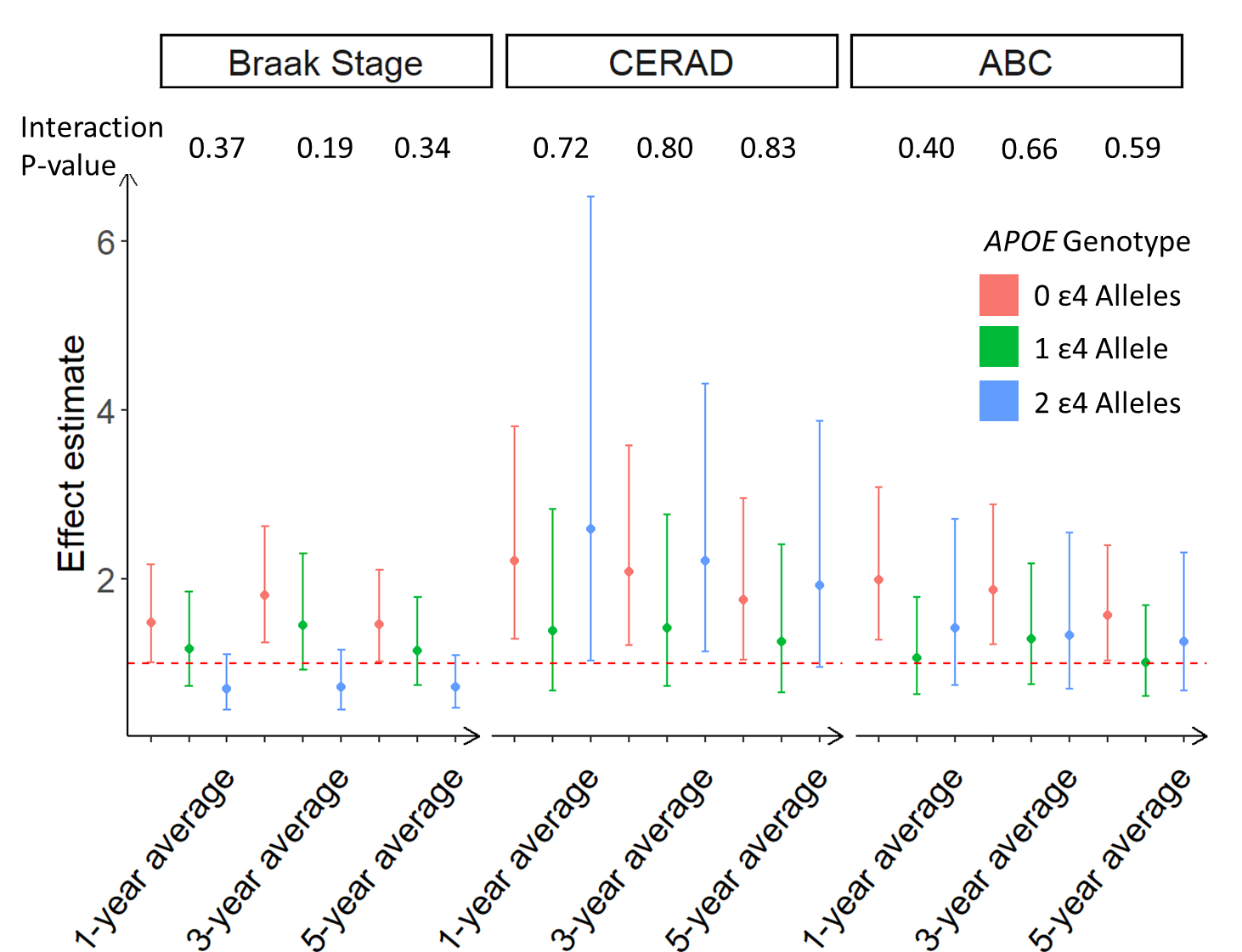

**Figure S6.** Association between PM_2.5_ and neuropathology markers stratified by *APOE* genotype (0, 1, 2 ε4 alleles). Associations were estimated using ordinal logistic regression, models adjusted for age at death, calendar year at death, race, sex, education, ADI, and a PM_2.5_**APOE* genotype interaction term.
